# *HSD3B1* genotype among metastatic castration-resistant prostate cancer patients: pharmacodynamics and clinical outcomes after treatment with abiraterone plus prednisone

**DOI:** 10.1101/2025.09.17.25335890

**Authors:** Diogo Assed Bastos, Denis Leonardo Fontes Jardim, Ricardo Zylberberg, Ernande Xavier dos Santos, Lilian Tiemi Inoue, Anamaria Aranha Camargo

## Abstract

The impact of *HSD3B1* (1245 A>C) polymorphism on clinical outcomes of patients with metastatic castration-resistant prostate cancer (mCRPC) treated with abiraterone remains under debate. We investigated how *HSD3B1* genotypes influence serum levels of testosterone, dehydroepiandrosterone sulfate (DHEA-S), abiraterone, delta-4-abiraterone (D4A) and clinical outcomes of mCRPC patients treated with abiraterone plus prednisone (AAP). Blood samples from 42 mCRPC patients were collected during AAP treatment. *HSD3B1* genotypes were determined by Sanger sequencing and analyses were made comparing AA versus AC+CC groups. PSA decline equal or above 50% from baseline (PSA50) at any time, Time to PSA progression (TPP), and Overall Survival (OS) were evaluated. Frequencies of AA, AC and CC *HSD3B1* genotypes were 50.0%, 33.3%, and 16.7%, respectively. The *HSD3B1* genotype did not influence steroids or drug levels during AAP treatment. PSA50 at any time was 71.4% for AA and 42.9% for the AC+CC group (p=0.061). Median TPP was similar between groups. The AA group had a median OS of 21.3 months, which was not reached by the AC+CC group (p=0.15). PSA50 at any time was lower in the AC+CC compared to AA group, though not statistically significant. The HSD3B1 genotype was not associated with TPP nor with OS.

## INTRODUCTION

Androgen deprivation therapy is the main systemic treatment for men with advanced prostate cancer (Sanda et al. 2018). However, treated patients ultimately develop castration-resistant prostate cancer (CRPC) despite castrated serum testosterone levels. Castration resistance in CRPC is driven by various mechanisms, including extragonadal androgen synthesis (Locke et al. 2008).

The protein 3β-hydroxysteroid dehydrogenase 1 (3β-HSD1), encoded by the *HSD3B1* gene, acts as a rate-limiting enzyme in the synthesis of extragonadal androgens (Simard et al. 2005). A common genetic polymorphism in the *HSD3B1* gene (1245 A>C) renders 3β-HSD1 less susceptible to proteasomal degradation, leading to increased protein half-life and higher intraprostatic androgen levels (Chang et al. 2013). Indeed, studies have revealed an association between *HSD3B1* (1245 A>C) polymorphism and poorer outcomes and shorter survival after castration in patients with castration-sensitive prostate cancer (CSPC) (Hearn et al., 2016, 2018; Agarwal et al. 2017; Shiota et al. 2019). Standard therapies for metastatic castration-resistant prostate cancer (mCRPC) include androgen-receptor pathway inhibitors, such as abiraterone and enzalutamide. In large phase III randomized trials, treatment with abiraterone and prednisone presented significant benefits for CRPC patients (de Bono et al. 2011; Fizazi et al. 2012; Ryan et al. 2013; Rathkopf et al. 2014). Abiraterone selectively and irreversibly inhibits CYP17A1, an enzyme crucial for androgen biosynthesis. Abiraterone is converted into multiple downstream metabolites by 3β-HSD1. One of the early metabolites, delta-4-abiraterone (D4A), strongly inhibits steroidogenic enzymes, such as CYP17A1 and 3β-HSD1, and suppresses androgen receptor signaling by directly binding to the androgen receptor (Li et al. 2015). However, further metabolites of D4A, including 3-keto-5-aplha-abiraterone, have pro-androgenic activity (Alyamani et al. 2018).

The *HSD3B1* (1245 A>C) polymorphism has been linked to increased generation of pro-androgenic metabolites (Alyamani et al. 2018). Therefore, even though abiraterone could be effective in blocking adrenal androgen synthesis, the increased conversion of abiraterone to pro-androgenic metabolites due to the *HSD3B1* polymorphism might reduce drug efficacy. Indeed, conflicting results have been reported regarding the association between *HSD3B1* genotype and clinical outcomes in mCRPC patients treated with abiraterone (Hahn et al. 2018; Khalaf et al. 2020; Lu et al. 2020).

Here, we investigated whether the *HSD3B1* genotype affected the serum levels of testosterone, dehydroepiandrosterone sulfate (DHEA-S), abiraterone, and D4A in mCRPC patients enrolled in the Abira-DES trial. These patients progressed after diethylstilbestrol (DES) and were treated with abiraterone acetate plus prednisone (AAP). Additionally, we investigated the association between the *HSD3B1* genotype and clinical outcomes in the Abira-DES trial. Genotypes were grouped to compare patients homozygous for the *HSD3B1* (1245A) allele with those carrying at least one *HSD3B1* (1245C) allele (AA versus AC+CC) to evaluate wether the presence of the *HSD3B1* (1245C) allele could impair abiraterone activity.

## MATERIALS AND METHODS

### Study design and participants

This was a retrospective, single-center, non-interventional study to explore *HSD3B1* genotype associations with testosterone, DHEA-S, abiraterone, and D4A serum levels using blood samples from 42 mCRPC patients enrolled in the phase II prospective, single-arm, Abira-DES study (NCT02217566). Participants in the Abira-DES study had progressed after diethylstilbestrol and received 1000mg abiraterone acetate orally in a fasting state, concomitantly with prednisone 5mg orally per day. All subjects remained on a stable regimen of androgen deprivation therapy (LHRH agonists) or were surgically castrated (bilateral orchiectomy). Peripheral blood samples were collected before initiating AAP therapy, after 12 weeks, and at the time of PSA progression according to the PCWG2 criteria. Data were collected between March 2020 and November 2020, including demographic information, previous treatments, Gleason score, PSA levels, time to PSA progression (TPP), and overall survival (OS). This study was approved by the Ethics Committee of Hospital Sírio-Libanês, São Paulo, Brazil (CAAE 24880719.5.0000.5461), and was performed in accordance with the Declaration of Helsinki. All patients provided their written informed consent to participate.

### Genetic and pharmacological analyses

*HSD3B1* genotype was determined by Sanger sequencing. A genomic fragment of approximately 200bp containing the *HSD3B1* (1245) region was amplified by PCR using the primers: 5’ GTCAAATAGCGTATTCACCTTCTCTTAT 3’ and 5’ GAGGGTGGAGCTTGATGACATCT 3’. Sequencing was performed in Seqstudio (ThermoFisher) and results were analyzed with Variant Reported software (ThermoFisher). Testosterone was quantified by liquid chromatography coupled with tandem mass spectrometry with a minimum detection limit of 9ng/dl. DHEA-S was quantified using a competitive electrochemiluminescence immunoassay with a minimum detection limit of 3μg/dl. Abiraterone and D4A were measured using ultra-performance liquid chromatography methodology coupled with mass spectrometry in Acquity I-Class/Xevo TQD. The detection methodology was developed at ApexScience laboratory, Campinas-SP, following standard recommendations (van Nuland et al. 2019). The minimum detection limit for abiraterone and D4A was 50pg/ml.

### Statistical analysis

The present study was a post-hoc analysis of blood samples collected during the Abira-DES study. No statistical calculation for power and sample size analysis was performed as the sample size was determined by the total number of participants with evaluable biological samples. Descriptive statistics were used to summarize the data. Genotypes were grouped to compare patients homozygous for the *HSD3B1* (1245A) allele with patients carrying at least one *HSD3B1* (1245C) allele (AA versus AC+CC). Patient baseline characteristics were compared using the Kruskall-Wallis test for continuous variables and the chi-square or exact Fisher test for categorical variables. Testosterone, DHEA-S, abiraterone, and D4A levels during AAP were compared using Student’s t-test. PSA50 at any time was evaluated using Pearson’s chi-square test and logistic regression model. The maximum PSA change at any time during AAP treatment was recorded using waterfall plots. TPP was determined as the difference between the participant’s inclusion date in the Abira-DES study and the PSA progression date according to PCWG2 criteria. OS was determined as the difference between the date of inclusion in the Abira-DES study and the date of death from any cause. TPP and OS were compared using the Kaplan-Meier method and the Log-rank test.

## RESULTS

### Patient population and *HSD3B1* genotype

We genotyped 42 patients enrolled in the Abira-DES study. The baseline clinicopathological characteristics of the cohort are listed in Table 1. The most common genotype was *HSD3B1* AA (50%, N=21), followed by AC (33.3%, N=14) and CC (16.7%, N=7).

**Table 1.** General characteristics of the study samples grouped by *HSD3B1* genotype.

|  |  | AA<br>(N=21) | AC+CC<br>(N=21) | TOTAL<br>(N=42) | p value |
| --- | --- | --- | --- | --- | --- |
| **Age at baseline**<br>**(years)** | N | 21 | 21 | 42 | 0.047 |
|  | Mean (SD) | 71.14 (7.29) | 66.29 (8.00) | 68.71 (7.95) |  |
| **Race, n (%)** | White / Caucasian | 16 (76.19) | 17 (80.95) | 33 (78.57) | 1 |
|  | Black or African/South American | 3 (14.29) | 4 (19.05) | 7 (16.67) |  |
|  | Asian/Pacific Islander | 1 (4.76) | 0 (0.00) | 1 (2.38) |  |
|  | Other | 1 (4.76) | 0 (0.00) | 1 (2.38) |  |
| **Gleason score, n (%)** | £6 | 2 (9.52) | 5 (23.81) | 7 (16.67) | <.0001 |
|  | 7 | 11 (52.38) | 4 (19.05) | 15 (35.71) |  |
|  | ³8 | 8 (38.10) | 10 (47.62) | 18 (42.86) |  |
|  | Unknown | 0 (0.00) | 2 (9.52) | 2 (4.76) |  |
| **Sites of Metastatic Disease<sup>a</sup>, n** | Visceral | 0 | 0 | 0 |  |
|  | Non Visceral | 28 | 26 | 54 |  |
|  | Bone | 19 | 18 | 37 |  |
|  | Node | 6 | 6 | 12 |  |
|  | Prostate mass | 2 | 2 | 4 |  |
|  | Soft tissue | 1 | 0 | 1 |  |
| **Prior radical prostatectomy, n (%)** | Yes | 2 (9.52) | 10 (47.62) | 12 (28.57) | 0.006 |
| **Prior radiotherapy to the prostate, n (%)** | Yes | 5 (23.81) | 10 (47.62) | 15 (35.71) | 0.1 |
| **Duration of prior DES therapy (months)** | N | 21 | 21 | 42 | 0.16 |
|  | Mean (SD) | 12.01 (11.33) | 11.97 (21.38) | 11.99 (16.90) |  |

<sup>a</sup> Some subjects presented more than one site of metastatic disease. n: number of subjects; SD: standard deviation.

We combined the AC and CC genotypes to compare them with the AA genotype in all analyses. The mean age at baseline was significantly higher in the AA group compared to AC+CC (71.1 versus 66.3 years, p=0.047). Gleason score ≥8 was more frequently observed in patients with AC+CC genotypes (47.6% versus 38.1%, p<0.001). Regarding prior radical prostatectomy, 9.5% of the AA group and 47.6% of the AC+CC group underwent surgery (p=0.006). Other characteristics, such as race, previous radiotherapy, and duration of prior DES therapy, did not differ between groups (Table 1).

### Pharmacodynamics and *HSD3B1* genotype

Testosterone and DHEA-S levels were measured at baseline, week 12, and at PSA progression. Abiraterone and D4A were measured at week 12 and at PSA progression. A total of 41 serum samples were available for dosage at baseline, 38 at week 12, and 36 at the time of PSA progression.

At baseline, all patients presented testosterone levels below the castration threshold (50ng/dl). Of these, 56.1% (23/41) had undetectable testosterone levels. Among the 18 patients with detectable levels, there was no statistically significant difference in the median testosterone levels between AA and AC+CC groups (14ng/dl (range 10.0-20.0 ng/dl) versus 12.5ng/dl (range 10.0-23.0ng/dl), respectively, p=0.989). By week 12, only one patient (2.6%) had detectable testosterone levels, and at PSA progression, all patients (36/36) had undetectable testosterone levels (Table 2).

**Table 2.**
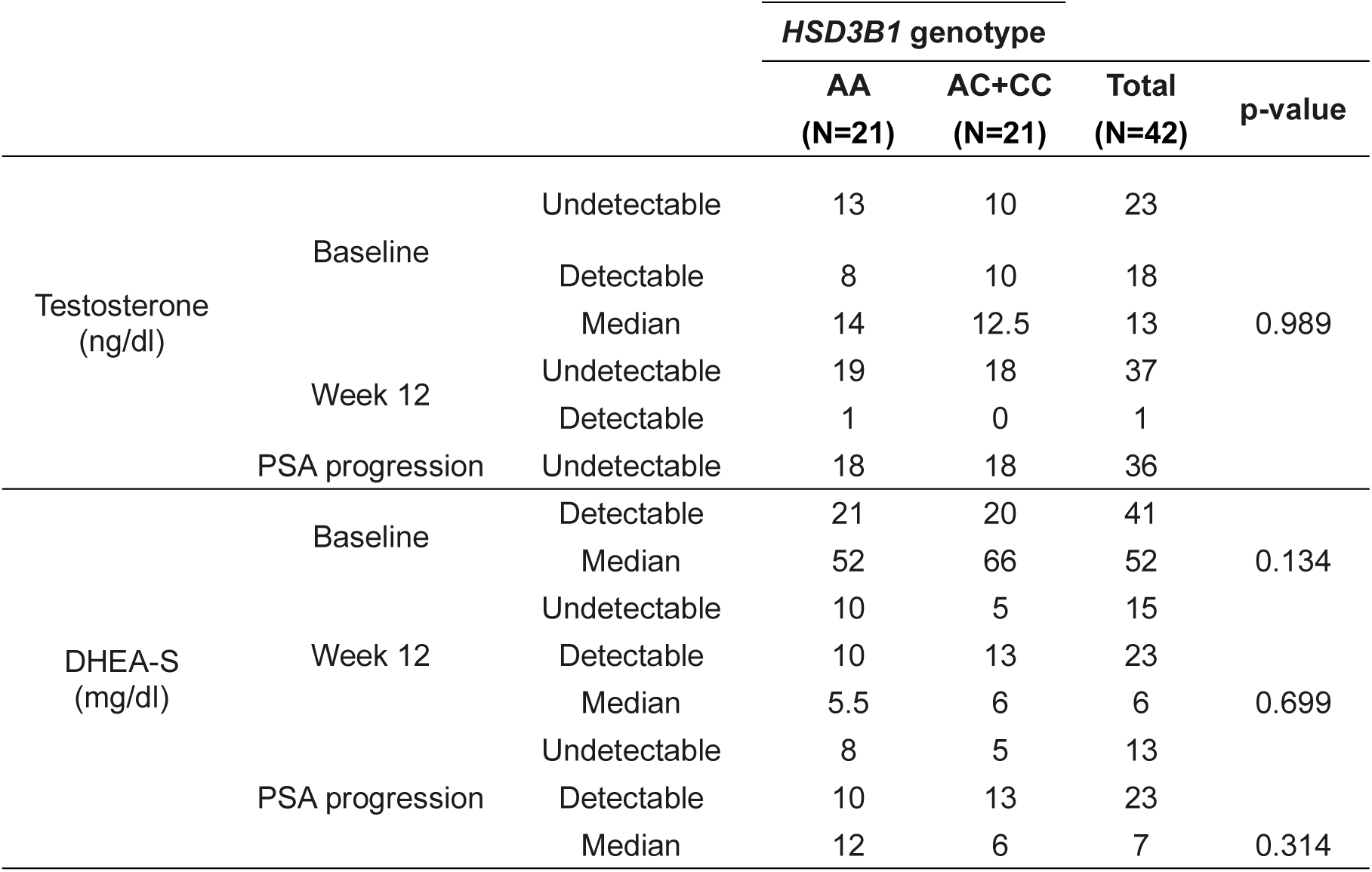
Testosterone and DHEA-S and *HSD3 B1* genotype.

All 41 patients had detectable DHEA-S levels at baseline, but there was no statistically significant difference in the median DHEA-S levels between AA and AC+CC groups (52.0μg/dl (range 6.0-178.0μg/dl) versus 66.0μg/dl (range 15.0-250.0μg/dl), respectively, p=0.134). By week 12, 60.5% (23/38) of the patients had detectable DHEA-S levels, but there was no significant difference in the median DHEA-S levels between groups (AA 5.5μg/dl (range 4.0-16.0μg/dl) versus AC+CC 6.0μg/dl (range 4.0-21.0μg/dl), p=0.699). At PSA progression, 63.9% (23/36) of the patients had detectable DHEA-S levels. Patients in the AA group had twice the median level of DHEA-S compared to the AC+CC group, however, the difference was not statistically different (12.0μg/dl (range 4.0-52.0μg/dl) versus 6.0μg/dl (range 4.0-27.0μg/dl), respectively, p=0.314) (Table 2).

Only one patient, at week 12, had undetectable abiraterone levels. There was no significant difference in the median abiraterone levels between AA and AC+CC groups (25.4ng/ml (range 4.5-367.0ng/ml) versus 36.7ng/ml (range 9.9-971.2ng/ml), respectively, p=0.267). At PSA progression, 11.1% (4/36) of the patients had undetectable abiraterone levels, and patients in the AC+CC group had approximately three times more abiraterone than the AA group; however, the difference was not statistically different (41.6ng/ml (range 0-825.0ng/ml) versus 13.9ng/ml (range 0.1-478.7ng/ml), respectively, p=0.322) (Table 3).

**Table 3.**
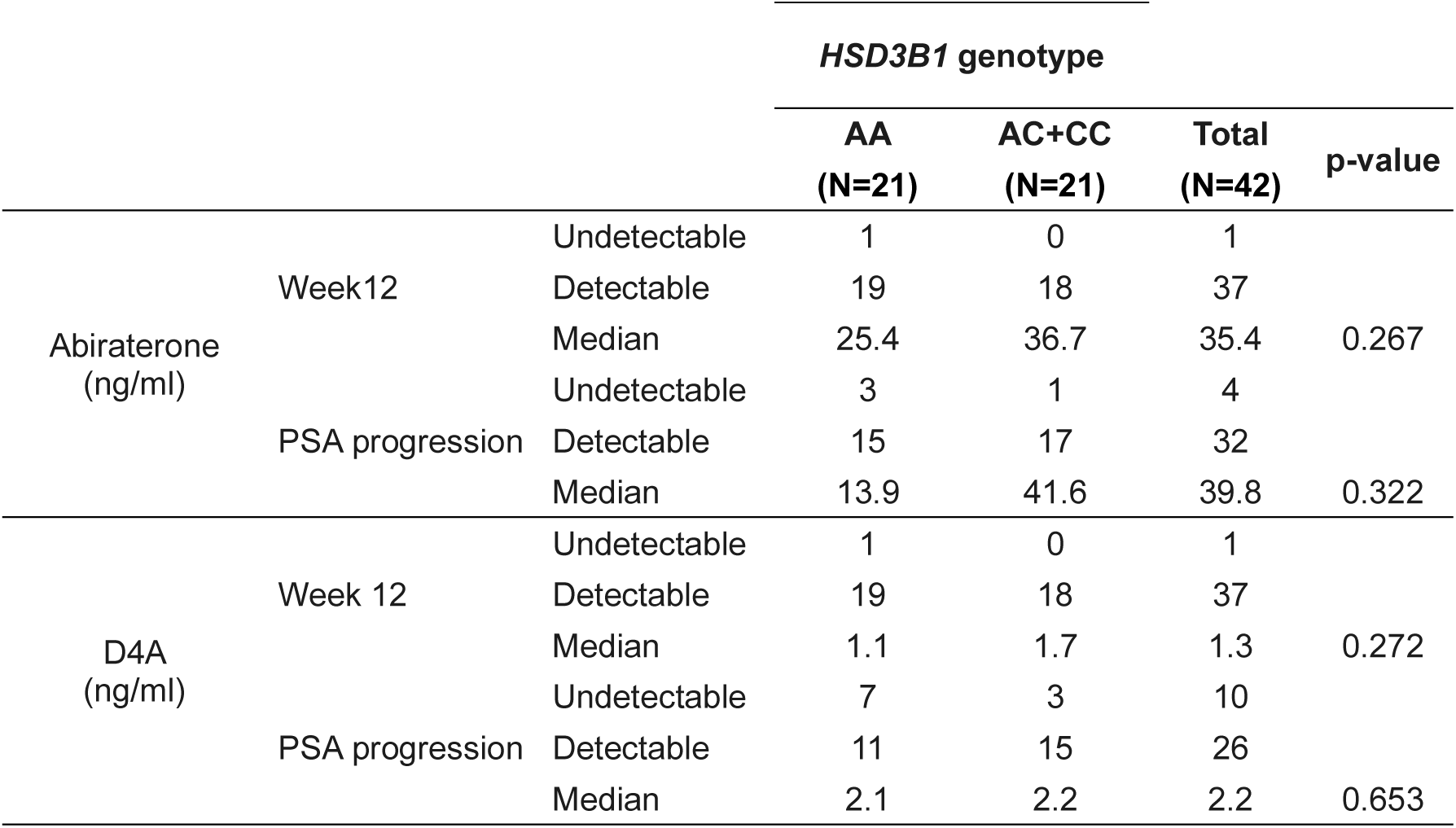
Abiraterone and D4A and *HSD3B1* genotype.

At week 12, one patient had undetectable D4A levels. There was no significant difference in the median D4A levels between AA and AC+CC groups (1.1ng/ml (range 0.2-6.8ng/ml) versus 1.7ng/ml (range 0.3-25.2ng/ml), respectively, p=0.272). At PSA progression, 27.8% (10/36) of them had undetectable D4A levels, and there was no significant difference in the median D4A levels between AA and AC+CC groups (2.1ng/ml (range 0.2-15.1ng/ml) versus 2.2ng/ml (range 0.2-15.3ng/ml), respectively, p=0.653) (Table 3).

### Clinical outcomes and *HSD3B1* genotype

PSA response was defined as a ≥ 50% decline in the PSA level (PSA50) at any time relative to the baseline PSA level. The PSA50 at any time was found to be 71.4% for AA group and 42.9% for AC+CC group (p=0.06), indicating a trend towards significance. We also observed a trend towards a higher chance of PSA50 at any time for the AA group (OR = 3.3 95%CI: 0.9-12.0, p=0.06). Maximum PSA change at any time was plotted and stratified by the *HSD3B1* genotype (Figure 1).

**Figure 1.**
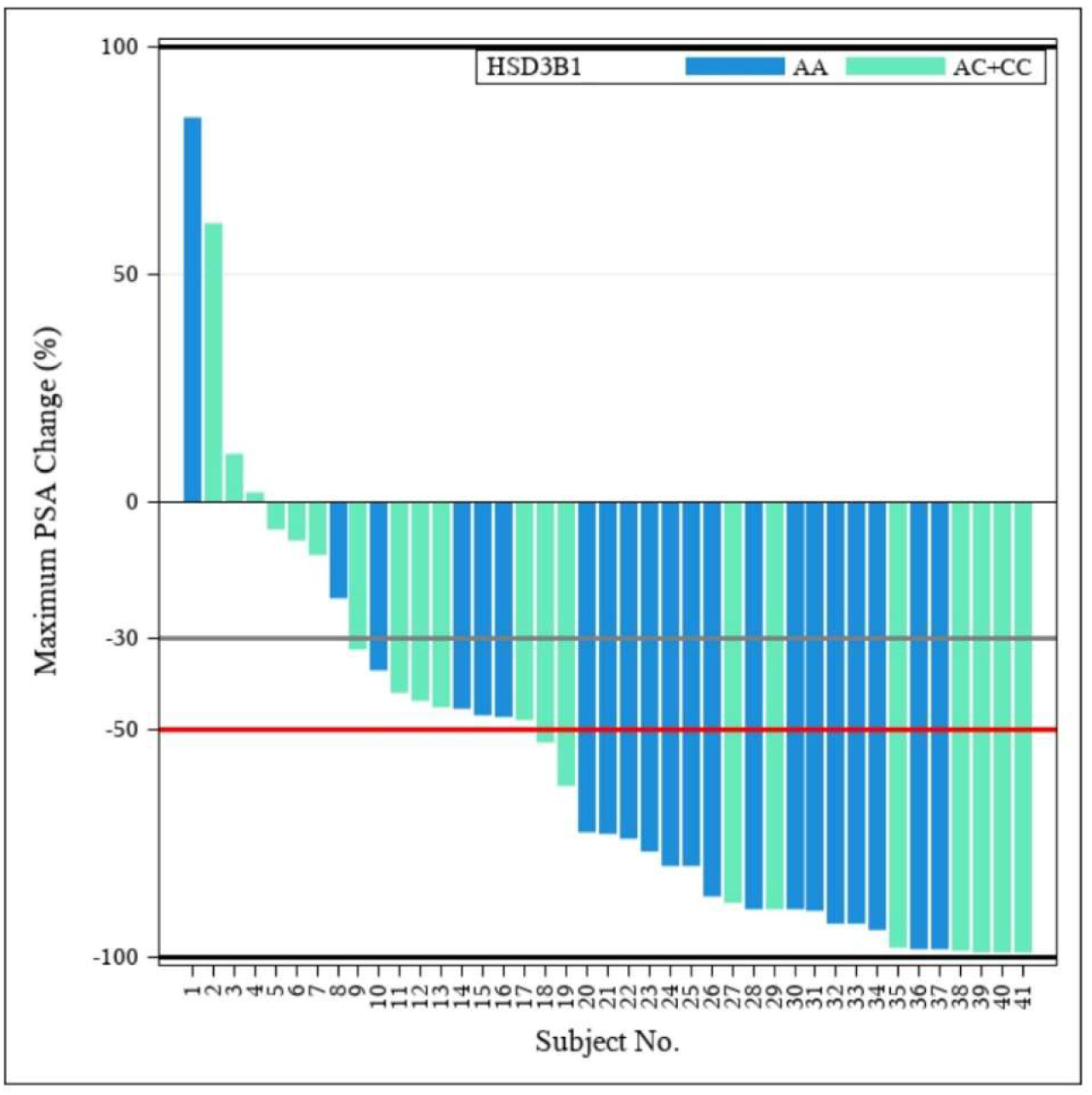
Waterfall plots displaying maximum PSA change at any time according to *HSD3B1* genotypes.

TPP was calculated as the difference between the date of participant inclusion in the Abira-DES study and the date of PSA progression according to PCWG2 criteria. The median TPP was 7.3 months (95%CI: 4.6-9.4; 10 events) for AA group and 7.9 months (95%CI: 5.6-14.4; 9 events) for AC+CC group (p=0.519) (Figure 2a).

**Figure 2.**
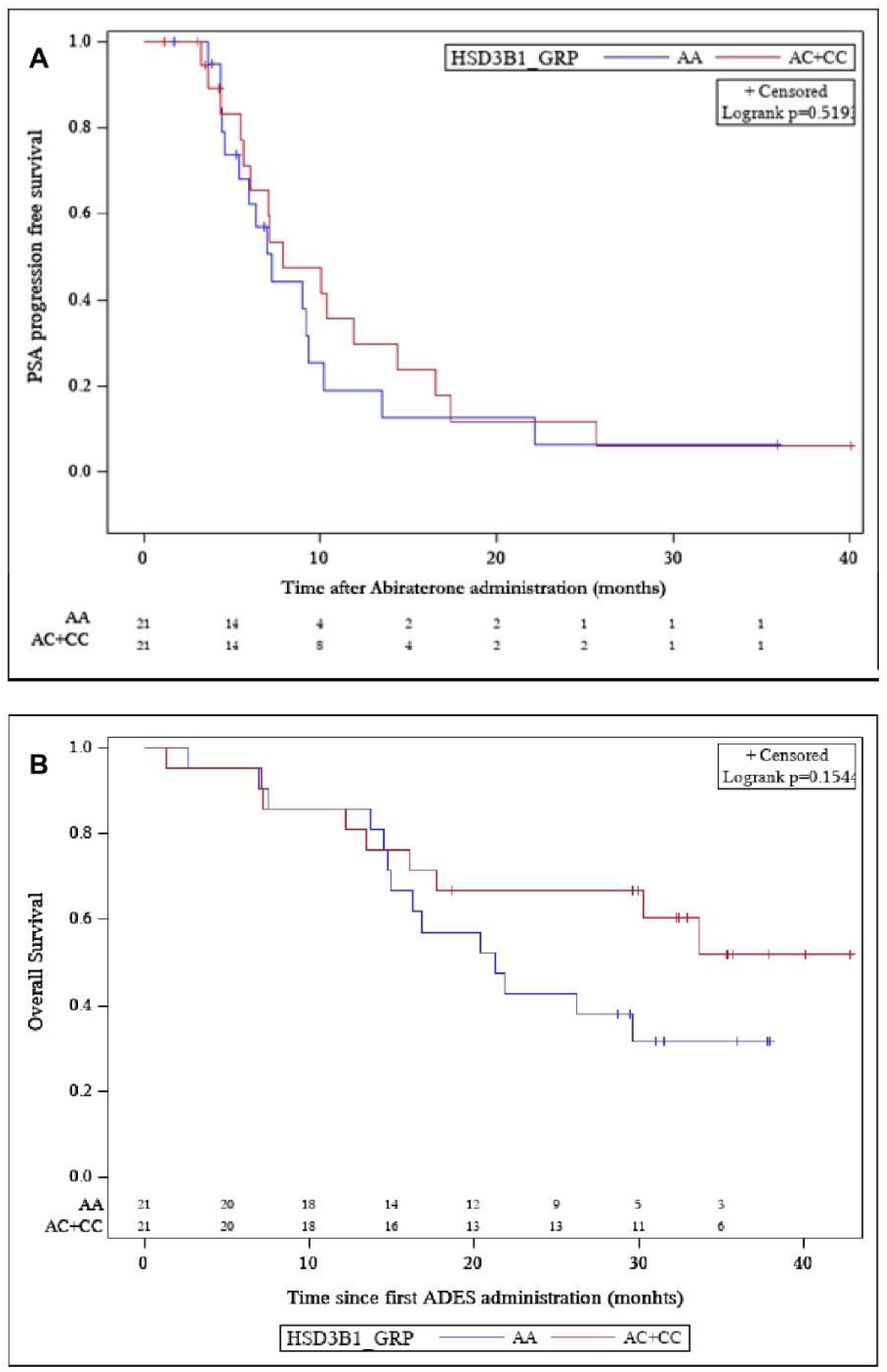
Kaplan-Meier estimates of the time (months) to (A) PSA progression and (B) overall survival according to *HSD3B1* genotypes.

OS was determined as the difference between the date of inclusion in the Abira-DES study and the date of death from any cause. Median OS was 21.3 months (95%CI lower limit: 14.8; 11 events) for AA group, which was not reached by AC+CC group (p=0.15) (Figure 2b).

## DISCUSSION

Our study was the first to report the prevalence of the *HSD3B1* (1245C) allele in a Latin American prostate cancer population. The frequencies of *HSD3B1*(1245) AA, AC, and CC genotypes were 50.0%, 33.3%, and 16.67%, respectively, which resemble previously reported data (Hahn et al. 2018; Almassi et al. 2018). Genotypes were grouped to compare patients homozygous for the *HSD3B1* (1245A) allele with patients carrying at least one *HSD3B1* (1245C) allele (AA versus AC+CC). Lower mean age at baseline and higher Gleason scores were more significantly observed in patients with AC+CC genotypes, suggesting a potential association between the *HSD3B1* (1245C) allele and a more aggressive disease.

No statistically significant differences were observed between the AA and AC+CC groups for testosterone or DHEA-S levels at baseline. Abiraterone and D4A serum levels were similar at week 12, independent of the *HSD3B1* genotype. Numerical differences in DHEA-S and abiraterone levels between AA and AC+CC groups were only observed at PSA progression. However, these results should be interpreted with caution due to the lack of control over the time between the last abiraterone dose and blood collection, as well as treatment adherence in the Abira-DES study.

The lack of significant differences in D4A serum levels between AA and AC+CC *HSD3B1* groups at week12 was unexpected, as individuals carrying the *HSD3B1* (1245C) allele were, in theory, anticipated to have higher D4A plasma levels compared to those carrying the wild-type allele. However, when analyzed individually, patients homozygous for the *HSD3B1* (1245C) variant allele had numerically higher D4A levels (1.9ng/ml) compared to patients homozygous for the *HSD3B1* (1245A) wild-type allele (1.1ng/ml) at week 12.

Accordingly, heterozygous patients had intermediary D4A levels (1.6ng/ml) at week 12. Also, investigating the effect of *HSD3B1* genotype impact on serum D4A levels in 19 CRPC patients, Shiota et al. (2023) observed higher D4A concentrations in patients heterozygous for *HSD3B1* allele (AC group) than in those homozygous for the *HSD3B1* (1245A) wild-type allele (AA group) after four weeks of treatment with abiraterone.

Remarkably, while most studies have focused on the relationship between the *HSD3B1* genotype and clinical parameters of mCRPC patients treated with CYP17A inhibitors (15-18,19), to the best of our knowledge, our study was the first to analyze these data alongside testosterone, DHEA-S, abiraterone, and D4A serum levels concomitantly and at different time points during AAP treatment.

In a study of 30 mCRPC patients receiving abiraterone, increased 3-keto-5α-abiraterone serum levels were associated with the presence of the *HSD3B1* (1245C) allele. However, no association between the *HSD3B1* genotype and PFS was observed (Alyamani et al. 2018). Another study of 68 patients with mCSPC or mCRPC treated with AAP also found no statistically significant association between the HSD3B1 genotype groups and plasma concentrations of abiraterone and D4A (Takahashi et al. 2023). Two other studies have investigated the link between abiraterone and derived metabolites plasma levels and the clinical outcomes of mCRPC patients treated with abiraterone, showing that those with lower abiraterone levels (van Nuland et al. 2020), or higher D4A levels and higher D4A/abiraterone ratio (Blanchet et al. 2018) had worse clinical outcomes. However, neither of these studies evaluated *HSD3B1* genotypes for any potential associations.

In our study, PSA50 at any time was numerically lower in the AC+CC group, suggesting that androgenic pathway inhibition was more effective in patients lacking the *HSD3B1* (1245C) allele. However, we found no significant association between *HSD3B1* genotypes and TPP or OS. Similar analyses of patients with advanced prostate cancer treated with CYP17A1 inhibitors have reported conflicting findings (Almassi et al. 2018; Hahn et al. 2018; Shiota et al. 2019; Khalaf et al. 2020; Lu et al. 2020). A study involving 90 mCRPC patients undergoing ketoconazole treatment showed that the *HSD3B1* (1245 A>C) polymorphism was associated with longer PFS, and the median therapy duration increased with the number of inherited *HSD3B1* (1245C) alleles (Almassi et al. 2018). Although ketoconazole is no longer routinely used in the clinic, the study indicated that mCRPC patients carrying the *HSD3B1* (1245C) allele were more responsive to the blockage of intratumoral androgen production. Similarly, in a cohort of 99 Japanese CRPC patients treated with abiraterone, the AC genotype group was associated with lower progression and mortality risks than the AA group (Shiota et al. 2019). On the other hand, a study involving 76 mCRPC patients treated with first-line abiraterone found no significant difference in PFS between AA and AC+CC *HSD3B1* genotype groups (Hahn et al. 2018). A two-cohort study with 547 mCRPC patients treated with first-line abiraterone or enzalutamide reported that men with *HSD3B1* CC genotype presented lower PSA50 and shorter time to progression. However, no clear association between *HSD3B1* genotype and clinical outcomes was found. In one of two cohorts that excluded prior docetaxel, OS was shorter, but not in the combined cohort, which included prior docetaxel (Khalaf et al. 2020). Additionally, in a two-cohort study with 266 mCRPC patients treated with first-line abiraterone or enzalutamide, the *HSD3B1* CC genotype was identified as an independent predictor of shorter OS (Lu et al. 2020).

The discrepancies between our study and the referenced studies could be partly attributed to variations in sample sizes, treatment approaches, racial disparities, and metabolic factors. Sharifi’s analysis of published data suggested poorer clinical outcomes for patients with the *HSD3B1* (1245C) allele after receiving abiraterone as initial treatment (Sharifi et al. 2020). Importantly, the mCRPC patients in our study were treated with DES prior to AAP. Additionally, genetic variations in *HSD3B1* may contribute to differences in treatment responses among racial groups. For instance, East Asian individuals have different prevalence of genetic variants and possibly distinct genomic risk factors compared to Caucasians (Fujimoto et al. 2013; Hearn et al. 2020). Notably, the study by Shiota et al. (2019), which reported superior clinical responses to abiraterone in CRPC men carrying the *HSD3B1*(1245C) allele, exclusively involved Japanese men, while other studies primarily focused on white/caucasian men, similar to our study (Khalaf et al. 2020; Lu et al 2020). Moreover, metabolic derivates, including abiraterone, D4A, and/or 3-keto-5α-abiraterone serum levels, may significantly influence clinical outcomes (Alyamani et al. 2018; Blanchet et al. 2018; van Nuland et al. 2020). Finally, when interpreting associations involving the *HSD3B1* gene, it is crucial to consider polymorphisms in other genes involved in the abiraterone metabolism. For instance, a study of 99 Japanese men with CRPC showed that analyzing polymorphisms in SRD5A2, which encodes an enzyme critical for the conversion of D4A to 3-keto-5α-abiraterone, in combination with *HSD3B1* genotype could enhance the prediction of abiraterone response (Shiota et al. 2021). In addition, CRPC patients homozygous for the *HSD3B1* (1245A) wild-type allele treated with abiraterone plus dutasteride, a 5α-reductase inhibitor, had longer time to treatment failure than heterozygous patients (Shiota et al. 2023).

Despite the novelty, our study has important limitations. Although patients had been treated under a prospective clinical trial, it was a non-randomized trial and included a small sample size, significantly limiting the statistical power of the study. Additionally, technical issues prevented measurements of 3-keto-5α-abiraterone serum levels, potentially impacting the accuracy of our results and underscoring the need for further research in this area. It is important to note, however, that this was a post hoc exploratory analysis of the Abira-DES trial, which was not originally developed to assess differences between *HSD3B1* genotype groups or to evaluate abiraterone pharmacodynamics or pharmacokinetics since the time between the last abiraterone dose and blood collection, as well as treatment adherence was not controlled. Therefore, further studies on clinical implications of *HSD3B1* genotype are needed to potentially guide treatment decisions. Briefly, we demonstrated that the *HSD3B1* (1245C) allele is present in approximately half of the Latin American prostate cancer population included in the Abira-DES study. The lower PSA50 at any time with abiraterone in the AC+CC group suggests possible pharmacodynamic implications of the *HSD3B1* genotype. Nonetheless, the lack of association of genotypes with steroids, abiraterone, and D4A levels during AAP treatment, as well as TTP and OS, implies that further studies are necessary.

## ACKNOWLEDGMENTS

We than statistician Rita Antonelli Cardoso for statistic analyses.

## DATA AVAILABILITY

Detailed data and materials used in this study may be available upon request.

## ETHICAL ISSUES

Written informed consent was obtained from all participants.

## AUTHORS CONTRIBUTIONS

DAB, DLFJ and AAC conceived the study. DAB, DLFJ, RZ and AAC were responsible for resources and supervision of the study; LTI and EXS conducted the genotyping experiments; DAB, DLFJ, LTI and AAC were responsible for investigation and writing original draft, review and editing. All authors read and approved the final manuscript.

## REFERENCES

1. Agarwal N, Hahn AW, Gill DM, Farnham JM, Poole AI, Cannon-Albright L (2017) Independent Validation of Effect of HSD3B1 Genotype on Response to Androgen-Deprivation Therapy in Prostate Cancer. JAMA Oncol 3:856–857.

2. Almassi N, Reichard C, Li J, Russell C, Perry J, Ryan CJ, Friedlander T, Sharifi N (2018) HSD3B1 and Response to a Nonsteroidal CYP17A1 Inhibitor in Castration-Resistant Prostate Cancer. JAMA Oncol 4:554–557.

3. Alyamani M, Emamekhoo H, Park S, Taylor J, Almassi N, Upadhyay S, Tyler A, Berk MP, Hu B, Hwang TH et al. (2018) HSD3B1(1245A>C) variant regulates dueling abiraterone metabolite effects in prostate cancer. J Clin Invest 128:3333–3340.

4. Blanchet B, Carton E, Alyamani M, Golmard L, Huillard O, Thomas-Scheomann A, Vidal M, Goldwasser F, Sharifi N, Alexandre J (2018) A PK/PD study of Delta-4 abiraterone metabolite in metastatic castration-resistant prostate cancer patients. Pharmacol Res 136:56–61.

5. Chang KH, Li R, Kuri B, Lotan Y, Roehrborn CG, Liu J, Vessella R, Nelson PS, Kapur P, Guo X et al. (2013) A gain-of-function mutation in DHT synthesis in castration-resistant prostate cancer. Cell 154:1074–1084.

6. de Bono JS, Logothetis CJ, Molina A, Fizazi K, North S, Chu L, Chi KN, Jones RJ, Goodman OB Jr, Saad F et al. (2011) Abiraterone and increased survival in metastatic prostate cancer. N Engl J Med 364:1995–2005.

7. Fizazi K, Scher HI, Molina A, Logothetis CJ, Chi KN, Jones RJ, Staffurth JN, North S, Vogelzang NJ, Saad F et al. (2012) Abiraterone acetate for treatment of metastatic castration-resistant prostate cancer: final overall survival analysis of the COU-AA-301 randomised, double-blind, placebo-controlled phase 3 study. Lancet Oncol 13:983–92.

8. Fujimoto N, Kubo T, Inatomi H, Bui HT, Shiota M, Sho T, Matsumoto T (2013) Polymorphisms of the androgen transporting gene SLCO2B1 may influence the castration resistance of prostate cancer and the racial differences in response to androgen deprivation. Prostate Cancer Prostatic Dis 16:336–340.

9. Hahn AW, Gill DM, Nussenzveig RH, Poole A, Farnham J, Cannon-Albright L, Agarwal N (2018) Germline Variant in HSD3B1 (1245 A > C) and Response to Abiraterone Acetate Plus Prednisone in Men With New-Onset Metastatic Castration-Resistant Prostate Cancer. Clin Genitourin Cancer 16:288–292.

10. Hearn JWD, AbuAli G, Reichard CA, Reddy CA, Magi-Galluzzi C, Chang KH, Carlson R, Rangel L, Reagan K, Davis BJ et al. (2016) HSD3B1 and resistance to androgen-deprivation therapy in prostate cancer: a retrospective, multicohort study. Lancet Oncol 171435–1444.

11. Hearn JWD, Xie W, Nakabayashi M, Almassi N, Reichard CA, Pomerantz M, Kantoff PW, Sharifi N (2018) Association of HSD3B1 Genotype With Response to Androgen-Deprivation Therapy for Biochemical Recurrence After Radiotherapy for Localized Prostate Cancer. JAMA Oncol 4:558–562.

12. Hearn JWD, Sweeney CJ, Almassi N, Reichard CA, Reddy CA, Li H, Hobbs B, Jarrard DF, Chen YH, Dreicer R et al. (2020) HSD3B1 Genotype and Clinical Outcomes in Metastatic Castration-Sensitive Prostate Cancer. JAMA Oncol 6:e196496.

13. Khalaf DJ, Aragón IM, Annala M, Lozano R, Taavitsainen S, Lorente D, Finch DL, Romero-Laorden N, Vergidis J, Cendón Y et al. (2020) HSD3B1 (1245A>C) germline variant and clinical outcomes in metastatic castration-resistant prostate cancer patients treated with abiraterone and enzalutamide: results from two prospective studies. Ann Oncol 31:1186–1197.

14. Li Z, Bishop AC, Alyamani M, Garcia JA, Dreicer R, Bunch D, Liu J, Upadhyay SK, Auchus RJ, Sharifi N (2015) Conversion of abiraterone to D4A drives anti-tumour activity in prostate cancer. Nature 523:347–351.

15. Locke JA, Guns ES, Lubik AA, Adomat HH, Hendy SC, Wood CA, Ettinger SL, Gleave ME, Nelson CC (2008) Androgen levels increase by intratumoral de novo steroidogenesis during progression of castration-resistant prostate cancer. Cancer Res 68:6407–6415.

16. Lu C, Terbuch A, Dolling D, Yu J, Wang H, Chen Y, Fountain J, Bertan C, Sharp A, Carreira S et al. (2020) Treatment with abiraterone and enzalutamide does not overcome poor outcome from metastatic castration-resistant prostate cancer in men with the germline homozygous HSD3B1 c.1245C genotype. Ann Oncol 31:1178–1185.

17. Rathkopf DE, Smith MR, de Bono JS, Logothetis CJ, Shore ND, de Souza P, Fizazi K, Mulders PF, Mainwaring P, Hainsworth JD et al. (2014) Updated interim efficacy analysis and long-term safety of abiraterone acetate in metastatic castration-resistant prostate cancer patients without prior chemotherapy (COU-AA-302). Eur Urol 66:815–825.

18. Ryan CJ, Smith MR, de Bono JS, Molina A, Logothetis CJ, de Souza P, Fizazi K, Mainwaring P, Piulats JM, Ng S et al. (2013) Abiraterone in metastatic prostate cancer without previous chemotherapy. N Engl J Med 368:138–148.

19. Sanda MG, Cadeddu JA, Kirkby E, Chen RC, Crispino T, Fontanarosa J, Freedland SJ, Greene K, Klotz LH, Makarov DV et al. (2018) Clinically Localized Prostate Cancer: AUA/ASTRO/SUO Guideline. Part I: Risk Stratification, Shared Decision Making, and Care Options. J Urol. 199:683–690.

20. Sharifi N (2020) Homozygous HSD3B1(1245C) inheritance and poor outcomes in metastatic castration-resistant prostate cancer with abiraterone or enzalutamide: what does it mean? Ann Oncol 31:1103–1105.

21. Shiota M, Narita S, Akamatsu S, Fujimoto N, Sumiyoshi T, Fujiwara M, Uchiumi T, Habuchi T, Ogawa O, Eto M (2019) Association of Missense Polymorphism in HSD3B1 With Outcomes Among Men With Prostate Cancer Treated With Androgen-Deprivation Therapy or Abiraterone. JAMA Netw Open 2e190115.

22. Shiota M, Akamatsu S, Narita S, Sumiyoshi T, Fujiwara M, Uchiumi T, Ogawa O, Habuchi T, Eto M (2021) The association between missense polymorphisms in SRD5A2 and HSD3B1 and treatment failure with abiraterone for castration-resistant prostate cancer. Pharmacogenomics J 21:440–445.

23. Shiota M, Inoue R, Tashiro K, Kobayashi K, Horiyama S, Kanji H, Eto M, Egawa S, Haginaka J, Matsuyama H (2023) A Phase II Trial of Abiraterone With Dutasteride for Second-Generation Antiandrogen- and Chemotherapy-Naïve Patients With Castration-Resistant Prostate Cancer. J Clin Pharmacol 63:445–454.

24. Simard J, Ricketts ML, Gingras S, Soucy P, Feltus FA, Melner MH (2005) Molecular biology of the 3beta-hydroxysteroid dehydrogenase/delta5-delta4 isomerase gene family. Endocr Rev 26:525–582.

25. Takahashi Y, Narita S, Shiota M, Miura M, Kagaya H, Kashima S, Yamamoto R, Nara T, Huang M, Numakura K et al. (2023) Impact of trough abiraterone level on adverse events in patients with prostate cancer treated with abiraterone acetate. Eur J Clin Pharmacol 79:89–98.

26. van Nuland M, Venekamp N, de Vries N, de Jong KAM, Rosing H, Beijnen JH (2019) Development and validation of na UPLC-MS/MS method for the therapeutic drug monitoring of oral anti-hormonal drugs in oncology. J Chromatogr B Analyt Technol Biomed Life Sci 1106–1107:26-34.

27. van Nuland M, Groenland SL, Bergman AM, Steeghs N, Rosing H, Venekamp N, Huitema ADR, Beijnen JH (2020) Exposure-response analyses of abiraterone and its metabolites in real-world patients with metastatic castration-resistant prostate cancer. Prostate Cancer Prostatic Dis 23:244–251.

